# The road to interruption of transmission of Gambiense Human African trypanosomiasis in the Democratic Republic of the Congo, an analysis of 25 years of routine data

**DOI:** 10.1101/2025.10.21.25338447

**Authors:** E Miaka, S Verschaeve, J Lebuki, T Mempongato Pongo Shampa, C Seghers, P Shih, R Inocencio da Luz, E Nicco, E Hasker

**Affiliations:** Programme Nationale de Lutte contre la Trypanosomiase Humaine Africaine (PNLTHA), Kinshasa, DRC; Institute of Tropical Medicine (ITM), Antwerp, Belgium

## Abstract

**Introduction:** Human African Trypanosomiasis (HAT) has caused hundreds of thousands of deaths in the Democratic Republic of the Congo (DRC) as recently as the 1990s. Since then annual incidence has reduced by 98% but the disease is known for its tendency of resurgence. We outlined areas at risk, HAT foci, and assessed trends in HAT infection within these foci.

**Methods:** We applied methodology developed by WHO to outline areas at risk by period and compared surface areas from 2000-2024. Combining risk areas identified we outlined HAT foci. Within each of these foci we assessed annual numbers of HAT cases and infection rates.

**Results:** Areas at risk have greatly reduced over the past 25 years, 67% for areas at low risk or above (≥ 1 per 100,000), 97% for areas at moderate to high risk (≥ 1 per 10,000). We outlined 15 HAT foci, all of which had been at moderate to high risk at some point in the 25-year period assessed. Three main foci account for over 80% of the total case load. Of those, the focus in the former province of Equateur rapidly declined in the early years of our analysis, while the foci in Kasai and Kwilu started to decline later. Currently 91% of all cases reported are within one of the foci outlined, most (77%) are clustered in the Kwilu focus.

**Discussion:** Great progress has been made on the road to interruption of HAT transmission in DRC. The epidemic is now concentrated mainly in a few foci. Given these results, the advent of a new single dose oral drug, acoziborole, makes it possible to consider interruption of transmission provided current efforts are sustained. Given the tendency of resurgence it is however imperative to put in place post-elimination surveillance in all foci.

## Introduction

Human African trypanosomiasis (HAT) is a chronic infectious disease transmitted by an insect vector, the tsetse fly. There are two forms of the disease, the acute east African variant caused by *trypanosoma brucei rhodesiense* (rHAT) and the chronic west African variant caused by *trypanosoma brucei gambiense* (*T*.*b. gambiense*, gHAT).^1 2^ Whereas rHAT is a zoonosis with wild and domestic animals as main reservoir host, gHAT is assumed to be anthroponotic. Historically 98% of all HAT cases reported have been gHAT.^3^ The disease is most common in remote rural areas and has a strongly clustered distribution in space and time. Areas at risk are often referred to as HAT foci.^4^

Over a period spanning more than two centuries, gHAT has caused devastating epidemics in west and central Africa.^5^ The latest epidemic, which heavily affected the Democratic Republic of the Congo (DRC), peaked in 1998 with almost 40,000 cases reported that year but estimates by WHO put the true prevalence about 10 times higher.^6^ Given that gHAT is fatal if left untreated and that no private sector treatment is available, the disease must have claimed hundreds of thousands of lives as recently as the 1990s. ^7 8^

In both forms of HAT, two disease stages are described. Stage 1 occurs soon after a person has been infected through the bite of a tsetse fly and presentation is mostly atypical with recurrent fever, headaches, adenopathy and pruritis as common symptoms.^1^ Once the parasite passes the blood-brain barrier, the patient reaches stage 2 which has more typical neurological symptoms, including behavioral changes, and will eventually lead to coma and death. It is assumed that all those infected will eventually progress to stage 2 and die, unless treatment is provided. Whereas rHAT is an acute disease in which patients rapidly progress to stage 2, for gHAT the duration of stage 1 and 2 are estimated at approximately 1½ years and 9 months respectively, unless the patient is treated.^9^

Given the chronic nature of gHAT and the fact that humans are assumed to be the only reservoir, gHAT control very much relies on active case finding and treatment.^10^ Active case finding by mobile screening teams became much more efficient when in the 1990s a serological screening test called card agglutination test for trypanosomiasis (CATT) was introduced.^11^ Populations living in the endemic foci are screened with CATT, those testing positive are subjected to parasitological confirmation. Parasitological confirmation relies on visualizing the parasite through microscopy, either a stained parasite or an unstained life parasite. The latter is much more sensitive but requires a concentration technique of which the mini anion exchange centrifugation for trypanosomiasis (mAECT) is most widely used.^12^ Until recently parasitologically confirmed cases still needed to undergo a lumbar puncture because treatment varied by disease stage.^13^

Until just over 15 years ago, treatment for stage 1 gHAT relied on intramuscular injections of pentamidine, whereas stage 2 was treated with melarsoprol, an extremely toxic arsenic derivative.^14^ This made it necessary, as described above, to perform a lumbar puncture to assess disease stage before starting treatment. Up to 10% of those treated with melarsoprol for stage 2 disease died from treatment even before they could die from the disease. A lot has changed in recent years. In 2009, melarsoprol was replaced by nifurtimox eflornithine combination therapy (NECT), greatly reducing adverse events.^15^ More recently an all oral drug, fexinidazole, was introduced, effective in both stages of the disease, obviating the need for the lumbar puncture unless the patient has signs of severe disease.^16 17^ Whereas fexinidazole still requires taking tablets for 10 days, a single dose oral drug, acoziborole, is now emerging from the development pipeline.^18^ Acoziborole appears to be highly effective in both stages of the disease. If safety can be confirmed also in parasitologically unconfirmed serological suspects, this would open the door to a ‘screen-and-treat’ approach. CATT positives or those testing positive to more recently introduced rapid tests for antibody detection can then be presumptively treated.^19^ Thus we might make diagnostic procedures a lot lighter and quicker and avoid losing cases because of imperfect sensitivity of confirmation tests, or simply due to non-availability of such tests. The fact that treatment is provided on the spot, without lengthy conformation procedures and without the possible need for a lumbar puncture might even increase uptake of screening. This is currently being evaluated in an ongoing trial in the north west of the DRC.^20^

The DRC has historically accounted for 60–70% of all reported gHAT cases. But from 2000 to 2024, annual case notifications declined from 16,975 to 330, representing a 98% reduction (https://www.who.int/data/gho/data/indicators/indicator-details/GHO/hat-tb-gambiense). The 2024 figure even includes 22 cases classified as ‘probable’, these are individuals in whom no parasites were visualized but who tested positive on more specific immunological and/or molecular tests at reference laboratory level. The country has a well-organized national HAT control program (PNLTHA) that provides both active and passive gHAT case finding and treatment, as well as vector control activities. HAT control activities in DRC have a long history, dating back to the colonial era. The PNLTHA have records available that go back as far as 1926. More detailed records are available since the year 2000 through the WHO HAT Atlas and very detailed records are available since 2019 through the PNLTHA TrypElim digital platform.^21^ The PNLTHA also produces annual reports with data on numbers screened and cases detected.

Continent wide we are now at a historic low, even less than the previous time in 1960. But this time we can go even further. Based on epidemiological data and availability of new tools, WHO have adopted interruption of transmission of *T*.*b. gambiense* by 2030 as a target in their NTD roadmap 2021-2030.^22^ In this effort DRC will obviously be a corner stone. In this analysis we will explore the vast array of data available to assess trends in gHAT risk in DRC over past 25 years at national and sub-national level. We aim to outline areas ever affected, the earlier mentioned foci, and within each of these foci we will asses progress towards interruption of transmission of *T*.*b. gambiense*.

## Methods

Data were obtained from three sources, the Trypelim database for the most recent data, starting from January 1^st^, 2019; the HAT Atlas database developed by WHO; and the annual reports of the PNLTHA for the period of 2000 till 2024. The HAT Atlas database contains aggregated data on active screenings conducted by location (village) and by year, as well as on patients diagnosed by location and by year, with a breakdown in those diagnosed through active and through passive case finding. The Trypelim database has individual records on patients diagnosed and treated, including location, age, gender, disease stage and treatment, as well as information on populations screened. The database starts in 2017, since January 1^st^, 2019 gHAT patients from all provinces of DRC have been included. Both databases have geographic coordinates for (almost) all villages included.

The primary focus of our analyses is the evolution of the area at risk for gHAT since January 1^st^, 2000. This is a key indicator defined by WHO in monitoring progress towards interruption of transmission of *T*.*b. gambiense*.^23^ We also explored trends in the proportion of participants found infected during active screening, at national and at sub-national levels for the same period, 2000-2024.

Areas at risk are calculated per year according to the methodology described by Simarro *et al*.^24^ Initially they used a 10-year period (2000-2009) but in their update for 2014 they used a 5-year window which they considered to strike a good balance between temporal resolution and robustness.^25^ Differentiation is made between five categories: 1) areas at very high risk (annual incidence ≥ 1 per 100), 2) areas at high risk (annual incidence < 1 per 100 and ≥ 1 per 1,000), 3) areas at moderate risk (annual incidence < 1 per 1,000 and ≥ 1 per 10,000), 4) areas at low risk (annual incidence < 1 per 10,000 and ≥ 1 per 100,000) and 5) areas at very low risk (annual incidence < 1 per 100,000). We applied the same methodology but on an annual basis using a rolling average, differentiating 21 overlapping 5-year periods from 2000-2004 until 2020-2024.

Our analysis was conducted in RStudio. We started from a data extract from HAT Atlas and Trypelim that has one line per year for each location in which HAT cases were reported or screenings conducted. Annual numbers of HAT cases aggregated at location level for each of the 21 overlapping 5-year periods mentioned above then were used to create heatmaps, applying a radius of 30 kilometer, a pixel size of one square kilometer, and with the number of cases per pixel as weight. Similarly we constructed population heatmaps based on Landscan images, for the same 5-year periods. Landscan raster images are available by calendar year, they have a resolution of one square kilometer, each pixel has a value that is an estimate of its population. For each period we overlaid the corresponding five Landscan raster images and obtained the sum of the population estimates for each pixel over the 5-year period. This raster image was then converted to points, each point having the value of the combined population estimate of the five pixels (years) it represents. This point layer was then transformed into a heatmap in the same way as described for the cases layer, with a diameter of 30 km, pixel size of one square kilometer and value of the point as weight. The case heatmaps were then overlaid with the corresponding population heatmaps and for each pixel the case layer was divided by the population layer. The resulting raster layer thus has a spatially smoothed incidence rate of HAT for each pixel. This raster layer was converted into two new raster layers, one in which we selected pixels with a value of ≥1 case per 10,000 population and one with pixel values ≥ 1 case per 100,000 population. These rasters were converted into shapefiles, dropping any polygon with zero value, indicating that it was below the incidence cutoffs specified. Next we calculated the surface areas in square kilometers, the sum of which is our area at moderate risk or above (for the layer of ≥ 1/10,000) or low risk or above (for the layer of ≥ 1/100,000). We present maps for the periods 2000-2004, 2005-2009, 2010-2014, 2015-2019 and 2020-2024 showing areas at moderate risk or above (categories 1-3 of WHO) and areas at low risk (category 4 of WHO). We also present a table showing annual the evolution in size of both types of areas, based on overlapping smoothed 5-year averages.

Another indicator that is used by the PNLTHA is the infection rate in active screening, calculated as the number of gHAT cases identified in active screening divided by the number of people screened. This is usually done per screening team or region and per year, for our analysis we have compiled the overall trend per year at national level. We fitted a Poisson model to determine annual proportions of change in infection rates. The model has been adjusted for probable inflection points. To identify these inflection points we applied a two-stage approach. As a first step we visually identified possible inflection points from the scatter plot between ‘year’ and ‘infection rate’, after which we employed a systematic grid search of piecewise linear regression models testing breakpoint combinations within the visually-identified ranges to determine the time points that minimized sum of squared residuals. We then created a new categorical variable ‘period’, each level combining the years between two inflection points (or before the first or after the last). We tested the ‘period’ variable and its interaction term in our Poisson model. As a final step we fitted separate models for each level of the ‘period’ variable to obtain the incidence rate ratio for each increment of one year within the period concerned.

We also compiled yearly infection rates per area at risk, the so-called HAT foci as explained in more detail below. These rates and their trends we just describe, without further statistical testing.

During a meeting convened by WHO in 2012, a total of 118 HAT foci were identified for DRC but without clear demarcations.^4^ For our analysis of annual infection rates in active screening at sub-national level we used HAT foci as target areas but to enhance operational feasibility, we redefined a smaller number of foci based on objective criteria. We plotted the earlier outlined areas at moderate risk or above (≥ 1 per 10,000) for successive 5-year periods i.e. 2000-2004, 2005-2009, 2010-2014, 2015-2019 and 2020-2024. These five layers we merged into a single layer, consisting of geographically separate polygons. From this layer we removed urban areas with populations above 150,000, based on shapefiles on ‘built up areas’ extracted from Grid3 (https://grid3.org/geospatial-data-drc) and population data from Landscan for 2023. We then extracted from the initial dataset with cases per location per year the total numbers of cases within each polygon over the entire 25-year period and dropped all polygons with less than 100 cases.

The remaining polygons we considered as our HAT foci and within each of those we recorded absolute numbers of cases (from active and passive case finding) and computed infection rates in active screening per year. Per focus and per year we extracted the total population screened in active screening and the number of HAT cases identified in active and in passive screening. We then calculated infection rates per focus by dividing numbers of cases identified in active screening by the population screened. Thus we aim to differentiate between foci at different levels of transmission.

As a final step we made estimates of populations to be kept under post-elimination surveillance in the HAT foci outlined. For this purpose we first identified all villages within each focus from which gHAT cases have ever been reported, either in active or in passive case detection. We then excluded villages from which less than five cases had ever been reported. As a next step we added population estimates for the villages retained, based on population estimates from the Trypelim database if available. For villages without available populations estimates we used 2023 Landscan data. For this purpose we constructed a common buffer of 1000 meters diameter around the centroid coordinates of each of the villages retained. Since the pixel size of Landscan is 1 km, this would (partially) cover 9 pixels, representing an area of 3 x 3 km for each village, unless if there would be spatial overlap between neighboring villages. We then merged all geographically continuous polygons and extracted population data from Landscan for each location retained.

This retrospective study was conducted with the approval and authorization of the PNLTHA. Ethics approval was obtained from the institutional review board of the Institute of Tropical Medicine, Antwerp, Belgium. (Ref. Nr. 1929/25). The requirement for informed consent was waived as the analysis involved only aggregated surveillance data that were fully anonymized, with no possibility of re-identification of individuals. The PNLTHA also authorized publication of the results of this analysis.

## Results

Altogether we found data on 141,251 HAT cases reported for the period 2000-2024. Out of those 137,662 (97.5%) had location data available. Among this group, 74,519 (54.1%) had been detected during 44,935,096 active screenings recorded, the remainder were detected through passive case finding. These represent 12,090 unique locations where there was ever a case. The annual numbers of locations from which a case was detected decreased from 6,548 in 2000 to 263 in 2024.

The area at risk rapidly decreased, starting from 2007, i.e. the period of 2005-2009. From 586,508 km^2^ at its height, the area at low risk and above decreased to 185,216 km^2^ for the period of 2020-2024, a 67% decrease. The area at moderate to high risk declined even faster from 335,091 km^2^ for the period 2005-2009 to 8,406 km^2^ for the period 2000-2024, a 97% reduction. Figure 1 below shows more of the details. Exact estimates detailing changes in area at risk are shown in annex 1.

**Figure 1:**
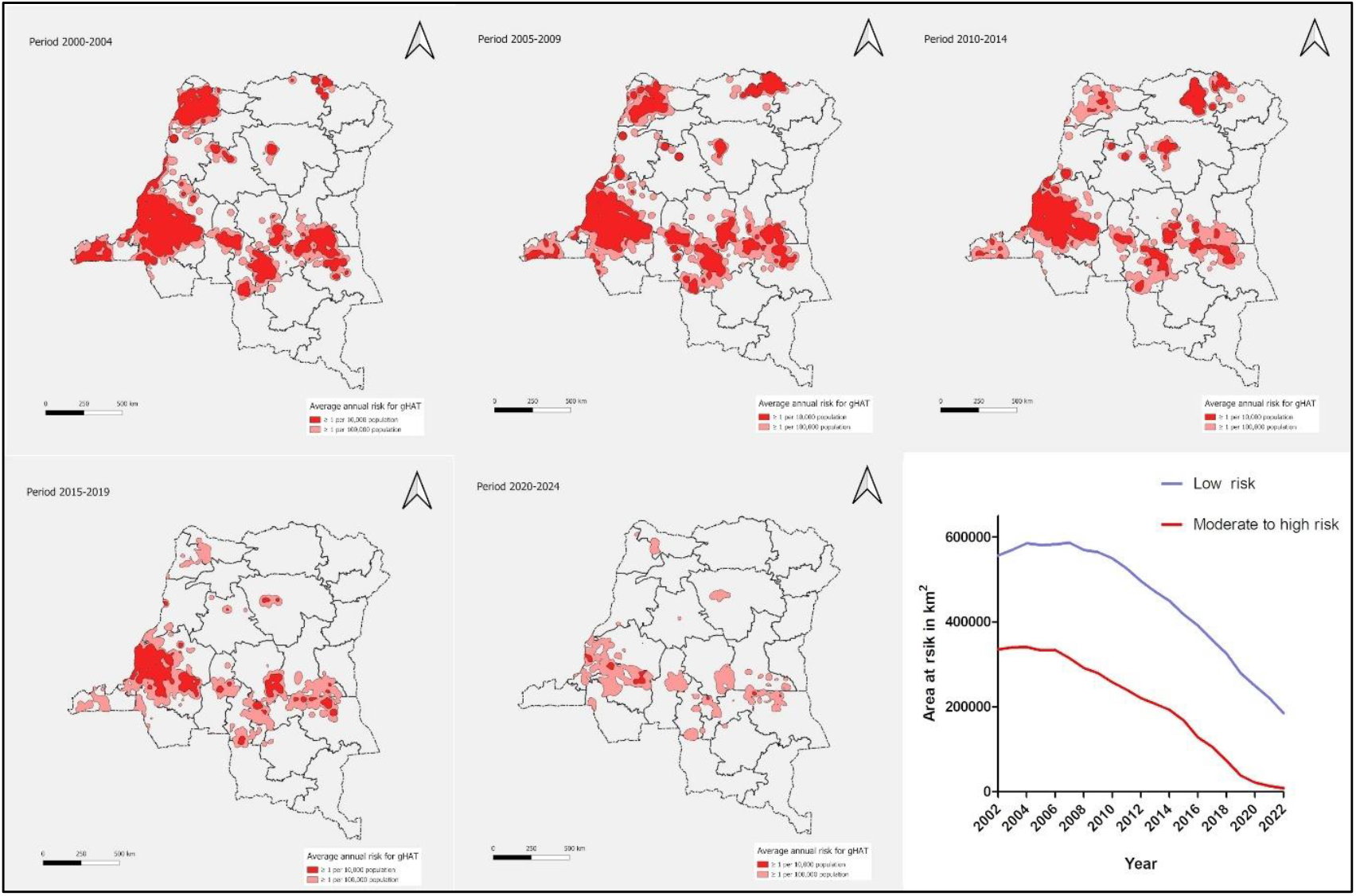
Areas at risk for HAT during the period of 2000-2024

When we plotted areas at moderate to high risk for the five successive 5-year periods from 2000-2004 until 2020-2024 and removed areas with less than 100 cases overall, we remained with 15 foci from which a total of 124,444 cases were reported over the 25-year period (90% of all cases with available location data). With the 2023 population estimates as denominator, cumulative incidence ranges from 8.2 to 85.2 per 10,000 population. Foci vary in size from 2,873 to 138,369 km^2^, current (2023) populations ranging from 38,835 to 7,654,948 (total population 22,533,215). Based on population estimates obtained for 4,324 endemic villages (i.e. villages with 5 or more cases over the period 2000-2024) in the 15 foci identified, the total population to be kept under post-elimination surveillance is approximately 7.5 million (7,492,635) of which 2.9 million are living in the Kwilu focus,1.7 million in the Ubangi focus and 1.2 million on the Kasai Oriental focus. The 15 foci identified are shown in figure 2 below, more details are provided in table 1.

**Table 1:**
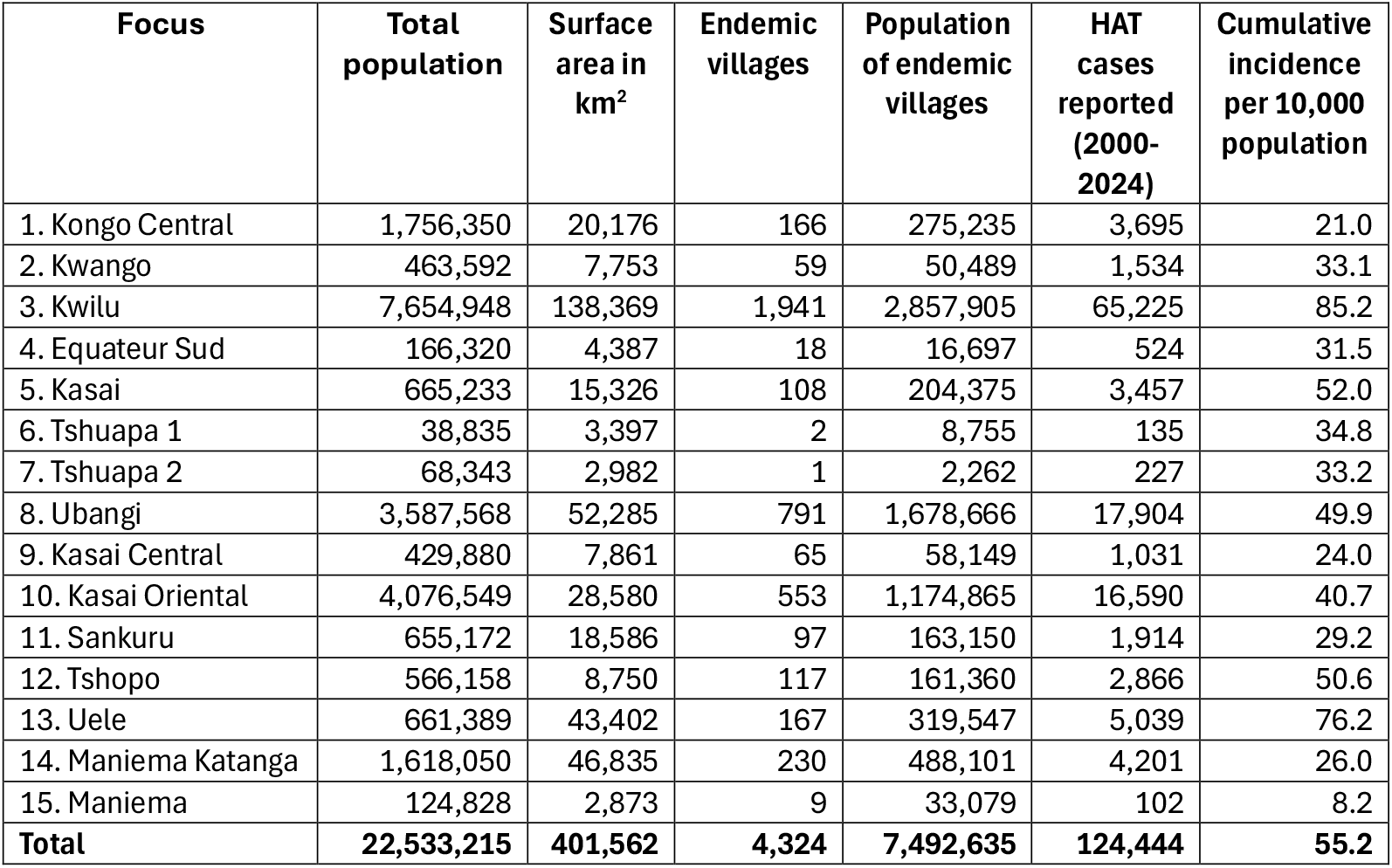
HAT foci outlined.

**Figure 2:**
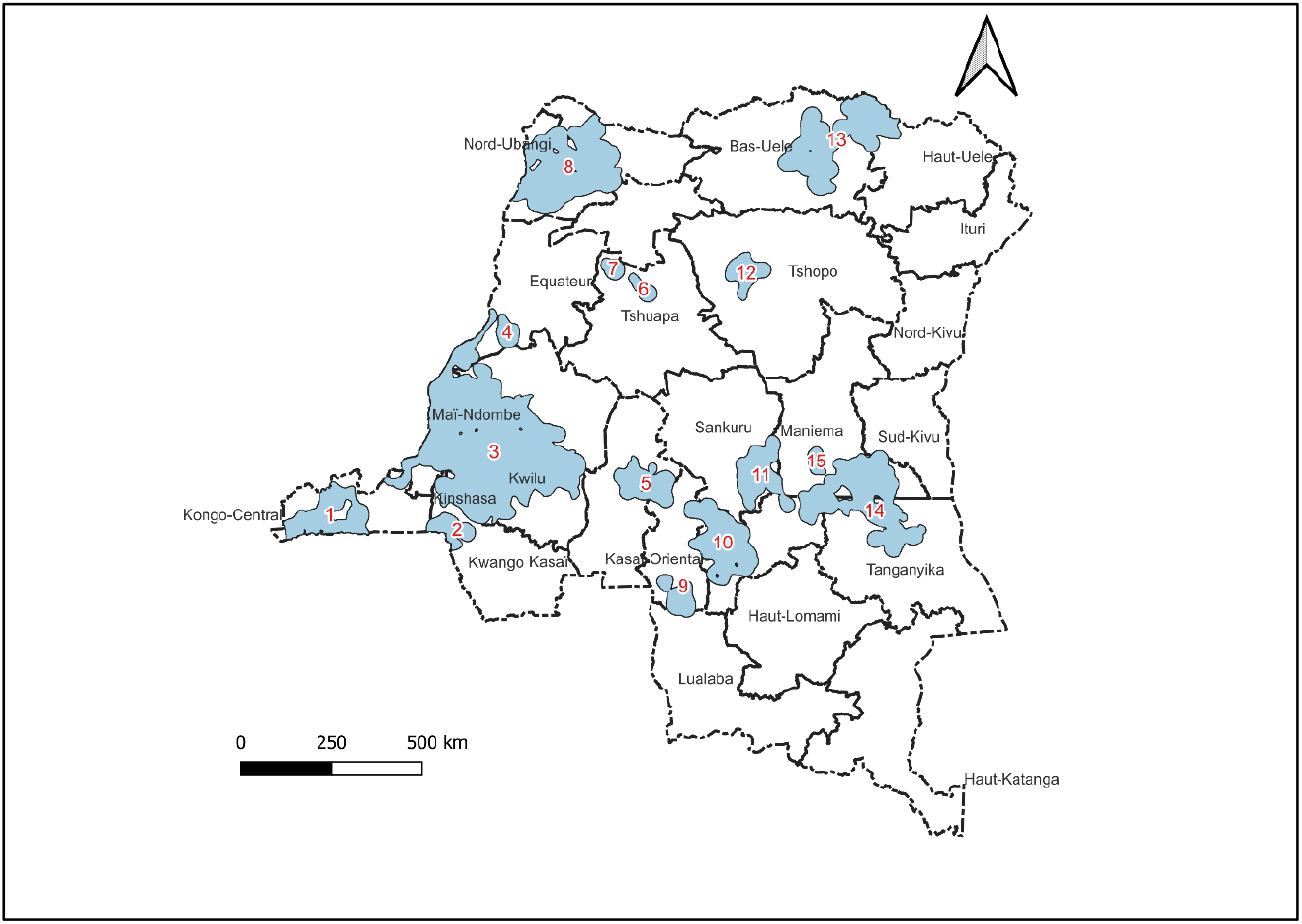
HAT foci outlined in DRC based on data for 2000-2024

Over the years, more than 80% of all cases notified are from the three main foci, Ubangi, Kwilu and Kasai Oriental. Out of 308 parasitologically confirmed cases reported in 2024, 237 (74%) were from the Kwilu focus, 40 (13%) were from the Kasai foci (mainly Kasai Oriental, 23 or 7.5%) and 41 (14%) were from the other foci. Altogether 280 out of 308 parasitologically confirmed cases reported in 2024 (91%), were from one of the foci identified. The remaining 28 were sporadic cases spread out across 10 provinces.

Trends in absolute numbers of cases, both actively and passively detected, are shown for the main foci in figure 3. There was a clear downward trend that started first in the Ubangi focus and was also the steepest there. Numbers of cases reported in the other foci came down more gradually.

**Figure 3:**
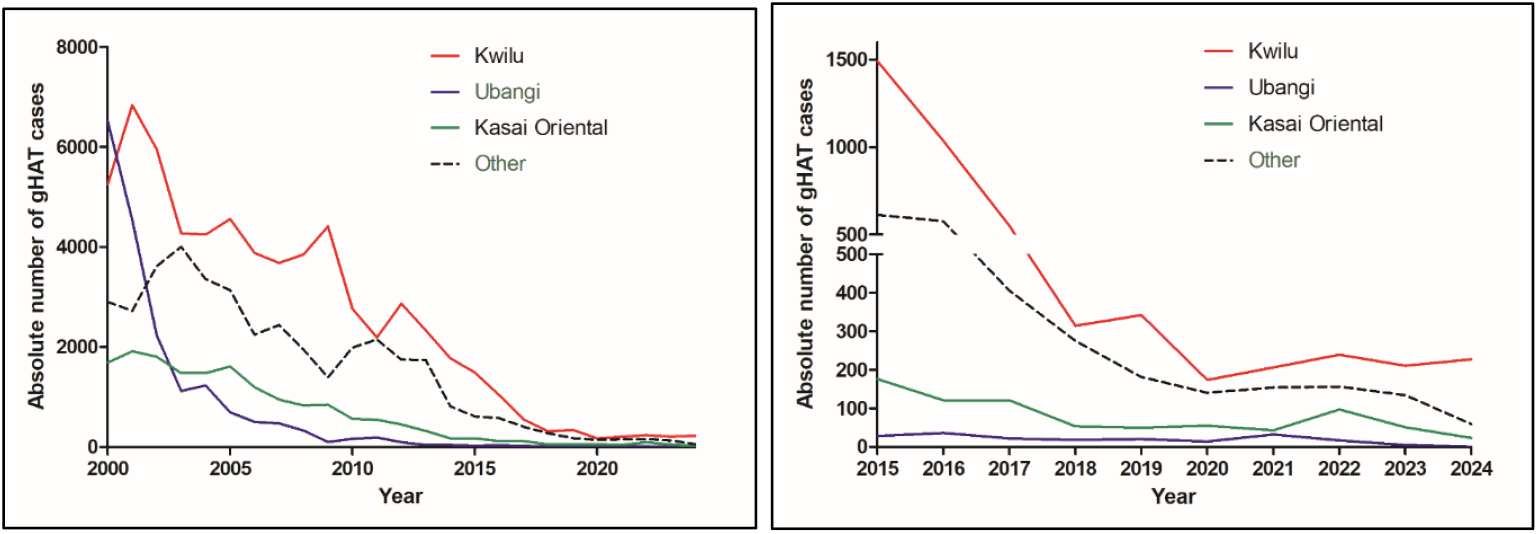
Evolution of absolute numbers of cases by focus, 2000-2025 (left panel), details for 2015-2024 (right panel)

At national level there was a strong and significant decrease in infection rate during active screening campaigns from above 60 per 10,000 in 2000 to less than 1 per 10,000 in 2024. Three periods could be discerned, from 2000-2004 there was a steep decline of 29.9% per year on average (95% CI 25.3-34.3%), from 2005-2010 there was a non-significant increase of 1.4% per year (95% CI −5.0 – 8.2%), and from 2011-2024 there was again a steep decrease of 23.5% per year (95% CI 18.9 – 27.9%). Figure 4 shows the trends, details are provided in annex 2.

**Figure 4:**
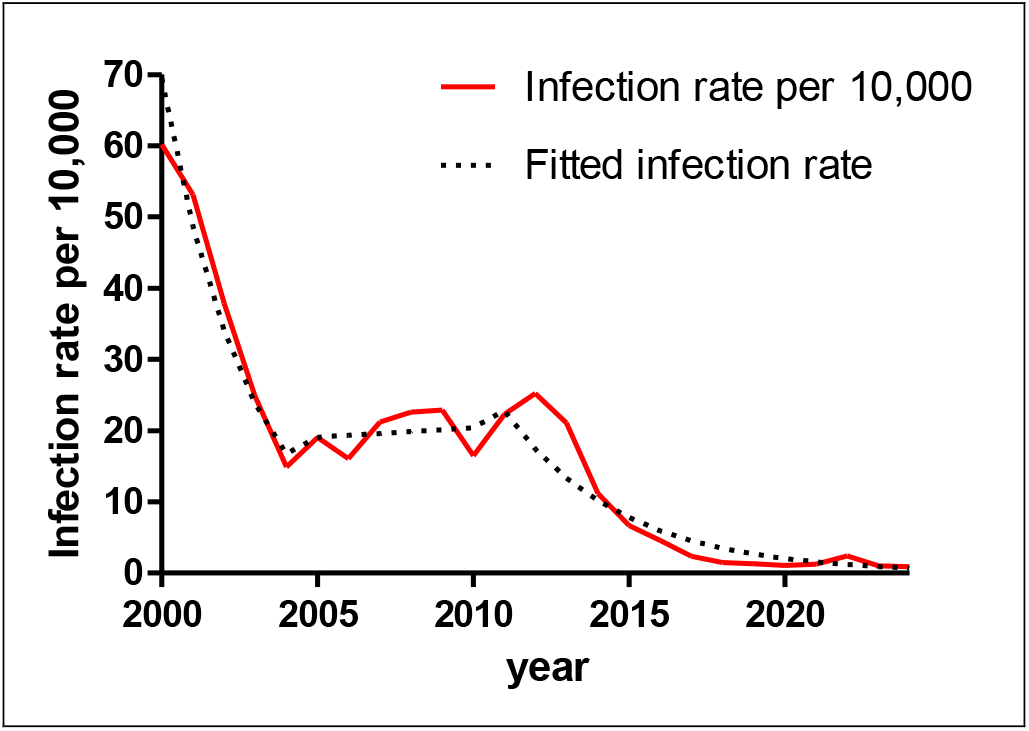
Infection rate with *T*.*b. gambiense* during active screening campaigns (2000-2024)

Overall the infection rate went down from 60.16 per 10,000 in 2000 to 0.93 per 10,000 in 2024, a 98% decrease. As shown in figure 5 below, at subnational level we found considerable differences in infection rates across the 15 foci identified. Infection rates have come down in all 15 foci and in 2024 were below 1 per 10,000 except for the Kwilu and the Kwango foci. Details on numbers screened and cases detected are presented in annex 3.

**Figure 5:**
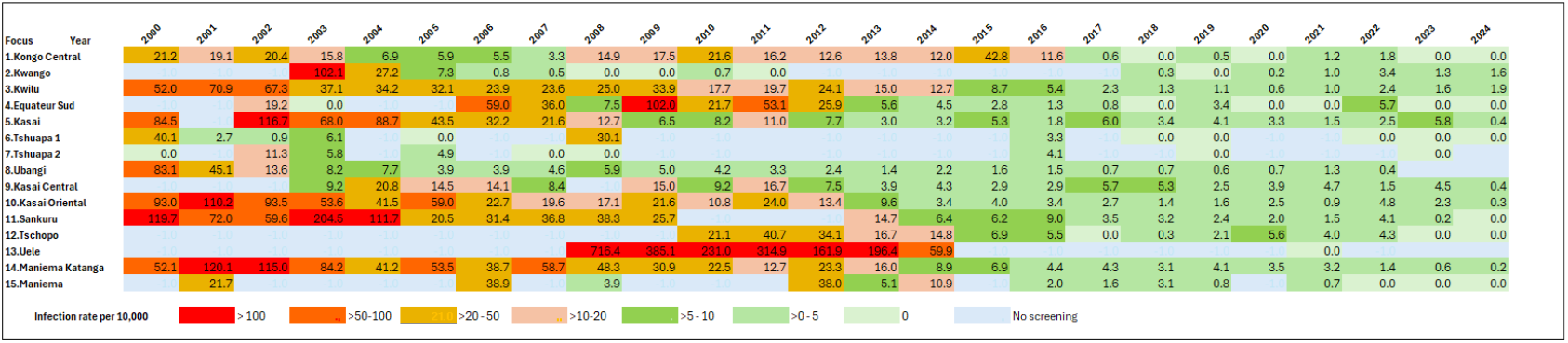
Infection risk during active screening campaigns by focus per year

## Discussion

Over the past 25 years enormous progress has been made on the road to interruption of transmission of *T*.*b. gambiense* in DRC. This is reflected not only in reduced numbers of gHAT cases notified but also in a strong reduction of area at risk and of infection risk among populations screened. The area at moderate to very high risk has been reduced by 97% since January 1^st^, 2000. At national level, infection risk among populations actively screened has reduced by 98% since then. The remaining hotspots are now concentrated in areas of the former Bandundu province, mainly Kwilu. In all gHAT foci identified, infection rates have also come down substantially or even reached zero but in Kwilu and Kwango they are still above of 1 per 10,000.

Elimination of transmission appears within reach but as numbers of cases decrease and large areas notify none or very few, identifying the last remaining cases is a challenge. It becomes imperative to maintain a strong quality assurance system, because with the current levels of case detection, inevitably diagnostic skills will deteriorate. This may either lead to false alarms or to true cases being missed. Already the PNLTHA has introduced video confirmation of microscopically confirmed cases.^26^ To assess whether cases have been missed due to imperfect sensitivity of classical confirmation tests and for quality assurance, samples are now collected of serological suspects for further screening at referral laboratory level. The algorithm is currently being optimized and will include one or more highly specific serological tests as well as a molecular test.

Another major challenge will be ensuring community participation in screening activities for a disease close to elimination and therefore no longer perceived as a priority. New strategies will need to be developed and tested, including combined surveillance with other neglected tropical diseases or with prevalent diseases such as malaria.

While a focus on currently active hotspots is clearly indicated, we also need to draw lessons from history. Based on the wealth of data available we are able to accurately outline the historic gHAT foci, most of which nowadays report very few cases. But these areas do require a well-designed mechanism of post-elimination surveillance that detects any resurgence at its earliest stage. Passive surveillance seems a logical option but may lack sensitivity as attendance rates at health facilities in rural areas of DRC are known to be very low.^27 28^ Post-elimination surveillance can make use of the tests at referral laboratory level described above, though dealing with prevalence levels below 1 per 10,000, imperfect specificity may become a major constraint.

Once the new drug, acoziborole, has been fully evaluated and safety confirmed in serological gHAT suspects as well, a screen & treat strategy might enable us to eliminate transmission also in foci that have so far remained active. An enormous reduction has been achieved over the past 25 years with tools that were far less performant than those currently available (or about to become available). The case finding and treatment algorithm used until now was assumed to miss up to 50% of prevalent gHAT cases.^10^ If this can be replaced by an easy to apply screen & treat strategy with immediate treatment on the spot for all serological gHAT suspects, elimination of transmission appears feasible, even in DRC. But having learned our lessons from the past, we know that post-elimination surveillance will be essential once transmission appears to have been interrupted. The total population to be kept under surveillance, currently estimated at 7.5 million, is manageable, given the fact that for the past 25 years close to two million per year on average have been actively screened. Contrary to active case finding, the purpose of post-elimination surveillance is to exclude the presence of HAT cases, not to find and treat them. For surveillance purposes it is therefore not necessary to screen each individual at risk, instead we can rely on inference drawn from samples. Optimal screening fractions and intervals now need to be worked out. This needs to take into account the time that has passed since elimination and the fact that (parts of) some foci may have undergone environmental changes, e.g. deforestation, that make resurgence less likely.

Though DRC has always contributed the largest numbers of gHAT cases, eliminating transmission in DRC alone will never be a lasting solution. During 2024, neighboring Central African Republic reported 111 reported cases and has now become the second largest contributor after DRC. It is imperative that successful gHAT control strategies are applied in all endemic countries.

### Limitations

This study has its limitations, it is based on surveillance data which are never fully comprehensive. Screenings are planned based on cases reported in preceding years but there may be areas that are left out altogether because no screenings have ever been conducted. Insecurity may also be a reason for not conducting screenings and areas thus automatically dropping off the list. As was pointed out, there may also have been resurgences in foci considered eliminated in which no further screening has been conducted for many years. Still the trends observed are very robust and indicative of interruption of transmission being possible.

## Conclusion

All available indicators show an enormous decrease in transmission of *T*.*b*.*gambiense*, making interruption of transmission a realistic prospect. Efforts will need to be sustained until a final mopping up campaign based on a screen & treat strategy with acoziborole is put in place continent wide. In the meantime we need to develop a reliable strategy for post-elimination surveillance which will be imperative for the foreseeable future but can be restricted to areas historically affected. Other than in 1960, we do now have the tools that should enable us to actually achieve interruption of transmission.

## Data Availability

All data produced in the present study are available upon reasonable request to the authors.

## Acknowledgements

We extend our sincere gratitude to the Gates Foundation (INV-030014 & INV-078633) and the Directorate-General for Development Cooperation of Belgium (Fifth Framework Agreement for Humanitarian Aid 2022–2026 DGD/ITM Program, BE-BCE_KBO-0410057701) for their generous funding support of HAT disease control programs and activities. Our deepest appreciation goes to the national sleeping sickness control programs, provincial health departments, and local health teams whose tireless efforts in disease surveillance and control, and whose commitment and expertise, made this work possible.

## Annex 1

**Table 1:** Area at risk over time (2000-2024)

| Period | low risk or above | moderate to high risk |  | Period | low risk or above | moderate to high risk |
| --- | --- | --- | --- | --- | --- | --- |
| 2000-2004 | 556274 | 335091 |  | 2011-2015 | 471322 | 207332 |
| 2001-2005 | 569627 | 340000 |  | 2012-2016 | 449560 | 192810 |
| 2002-2006 | 584783 | 340730 |  | 2013-2017 | 417626 | 167975 |
| 2003-2007 | 580775 | 332671 |  | 2014-2018 | 391271 | 128047 |
| 2004-2008 | 582167 | 333574 |  | 2015-2019 | 357064 | 105576 |
| 2005-2009 | 586508 | 314577 |  | 2016-2020 | 325323 | 73172 |
| 2006-2010 | 569158 | 291265 |  | 2017-2021 | 279129 | 38253 |
| 2007-2011 | 564007 | 279184 |  | 2018-2022 | 249241 | 21670 |
| 2008-2012 | 549423 | 257868 |  | 2019-2023 | 221021 | 13441 |
| 2009-2013 | 525872 | 240355 |  | 2020-2024 | 185216 | 8406 |
| 2010-2014 | 496298 | 220414 |  |  |  |  |

## Annex 2

Trends in infection risk at national level

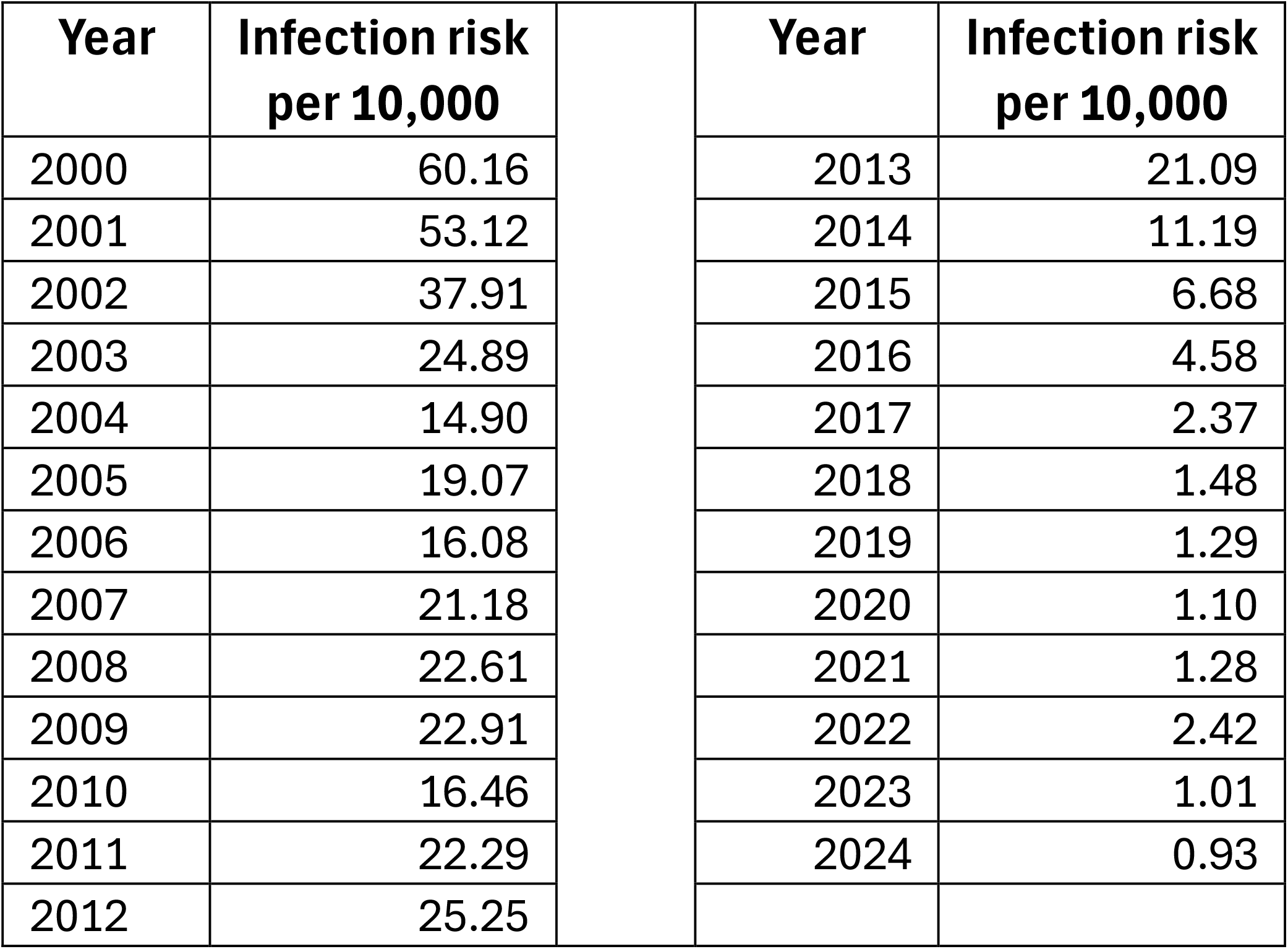

## Annex 3

Screening data

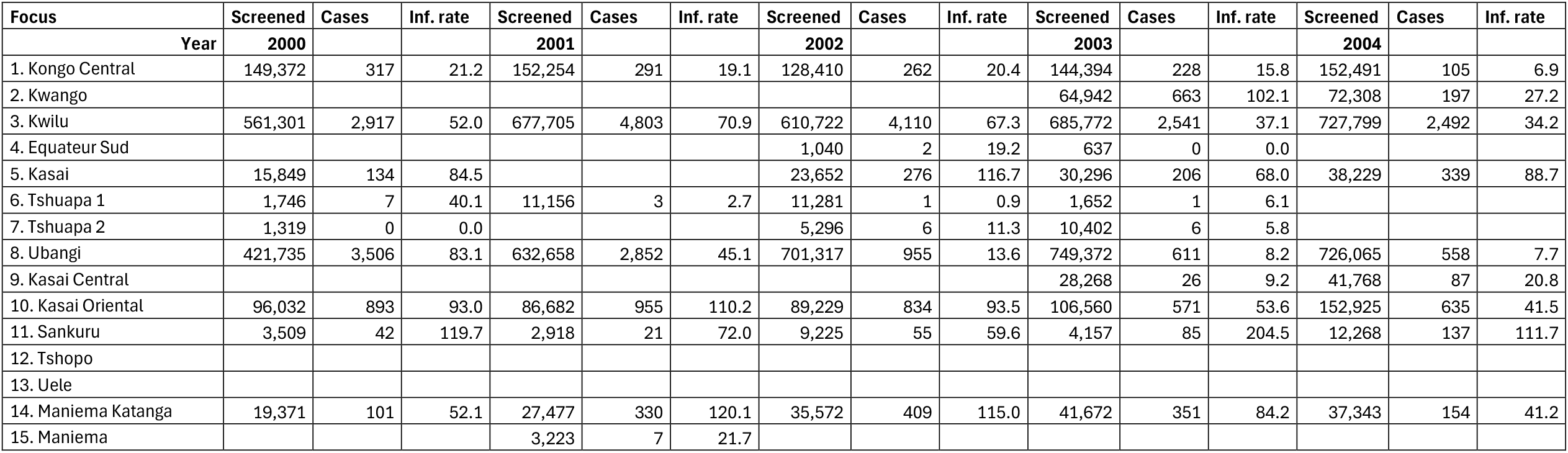
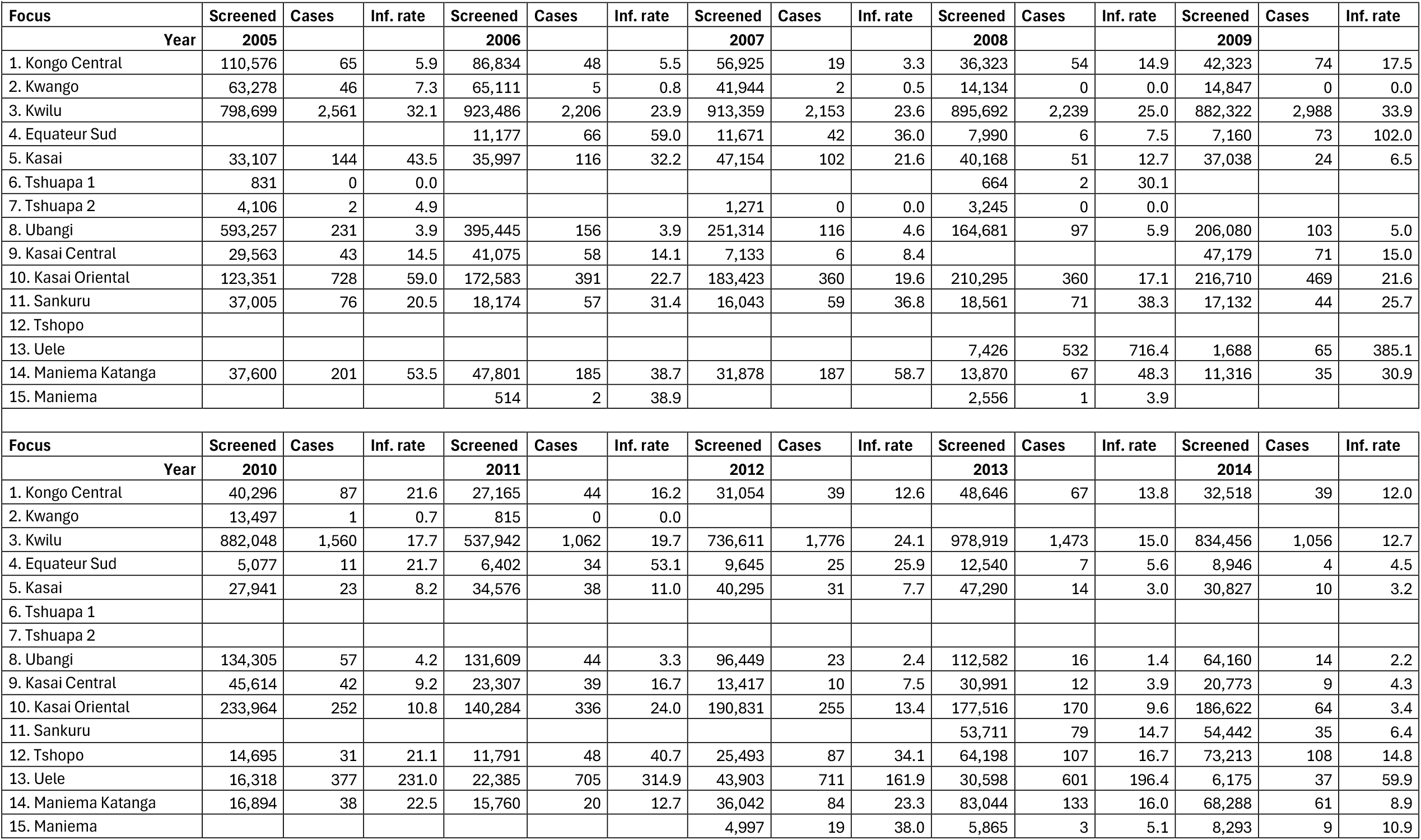
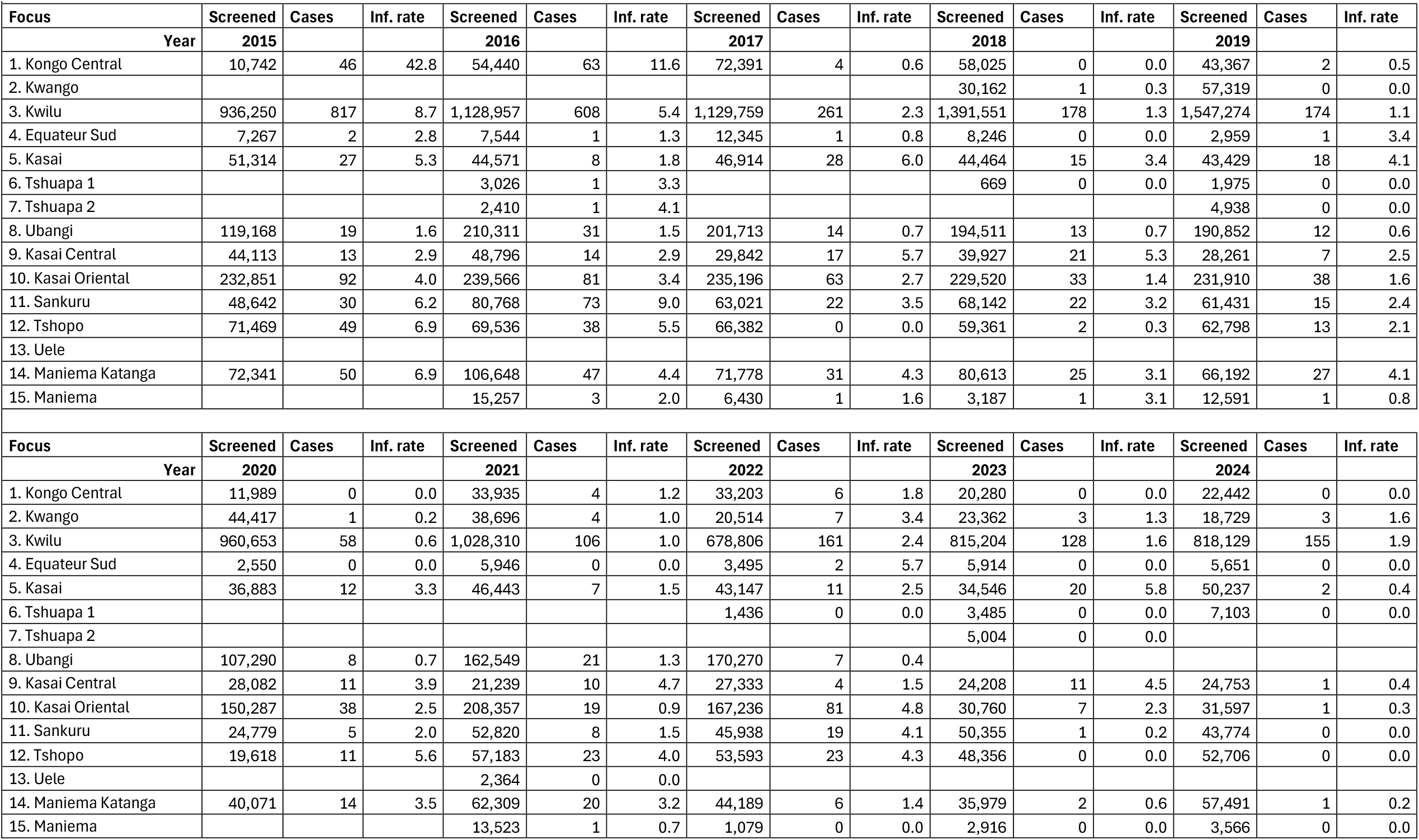

